# Country Level Incidence of Alzheimer Disease and Related Dementias is Associated with Increased Omega6 PUFA Consumption

**DOI:** 10.1101/2024.08.07.24311637

**Authors:** Timothy H. Ciesielski, Giuseppe Tosto, Razaq O. Durodoye, Farid Rajabali, Rufus O. Akinyemi, Goldie S. Byrd, William S. Bush, Brian W. Kunkle, Christiane Reitz, Jeffery M. Vance, Margaret A. Pericak-Vance, Jonathan L. Haines, Scott M. Williams

## Abstract

**INTRODUCTION:** Clinical and genetic studies have implicated lipid dysfunction in Alzheimer Disease (AD) pathogenesis. However, lipid consumption at the individual-level does not vary greatly within most cohorts, and multiple lipids are rarely measured in any one study.

**METHODS:** Mean country-level lipid intakes were compared to Age-Standardized Alzheimer-Disease-Incidence-Rates(ASAIR) in 183 countries across all inhabited continents. Penalized spline regression and multivariable-adjusted linear regression, including a lag between intake and incidence, were used to assess the relationships between five lipid intakes and ASAIR. Validation was conducted using longitudinal within-country changes between 1990 and 2019.

**RESULTS:** Omega6 Polyunsaturated-Fatty-Acid(PUFA) intake exhibited a positive linear relationship with ASAIR(multivariable-adjusted model: β=2.44; 95%CI: 1.70, 3.19; p=1.38×10^-9^). ASAIR also increased with saturated-fat, trans-fat, and dietary-cholesterol up to a threshold. The association between Omega6-PUFA and ASAIR was confirmed using longitudinal intake changes.

**DISCUSSION:** Decreasing Omega6-PUFA consumption on the country-level may have substantial benefits in reducing the country-level burden of AD.

## Introduction

Convergent evidence from genetics, neuropathology, laboratory experiments, and epidemiology indicates that lipids play a central role in the development of Alzheimer Disease (AD).^1–9^ However, the relative importance of different lipids remains unclear, and individual-level epidemiologic studies are often not well suited to address this question. Large cohort studies rarely have precise dietary information on specific lipid intakes over decades, and even where these data are available, the lipid intakes among individuals do not typically vary enough within a study population to discern patterns of association. Additionally, multiple categories of lipids are not usually available in one dataset, so their relative associations cannot be compared. Lastly, the potential for nonlinear relationships is rarely considered. These caveats often lead to equivocal conclusions upon meta-analysis.^10^ However, each of these issues is addressable with ecological analyses^11^ on the country-level. Mean intake levels for several lipid categories have now been measured with precision and they vary substantially among nations.^12,13^ The range and density of these data should allow for: 1) resolution of associations with AD incidence, and 2) the use of free-knot penalized spline regression^14^ to assess for nonlinear relationships. Together these factors provide strong impetus for conducting ecological analyses on the country-level that can characterize the relationship between large-scale lipid intake variations and AD.

Here we obtained country-level data on five lipid intakes^12,13^, and compared them to age-standardized Alzheimer Disease Incidence rates (ASAIR)^15^ using free-knot penalized spline regression and linear regression. We then attempted to validate our findings using longitudinal analyses that compared changes in country-level lipid intakes over 30 years to changes in ASAIR. These analyses do not seek to make inferences about individual level intakes and the risk of Alzheimer Disease for individuals. Thus, the ecological fallacy is not a relevant concern as both the exposure and the outcome are on the country-level. Country-level dietary data is the most appropriate data for studying country-level incidence rates and country-level interventions.

## Results

The mean ASAIR among the 183 countries was: 91.6 new cases per 100,000 per year with a standard deviation of 11.7 (Table 1). Mean country-level Omega 6 PUFA, saturated fat, and trans-fat intakes were 5.1%, 11.6%, and 1.1% of total energy intake, respectively, and the mean intakes for cholesterol and Omega3 PUFA were 323 and 249 mg/day, respectively.

**Table 1:**
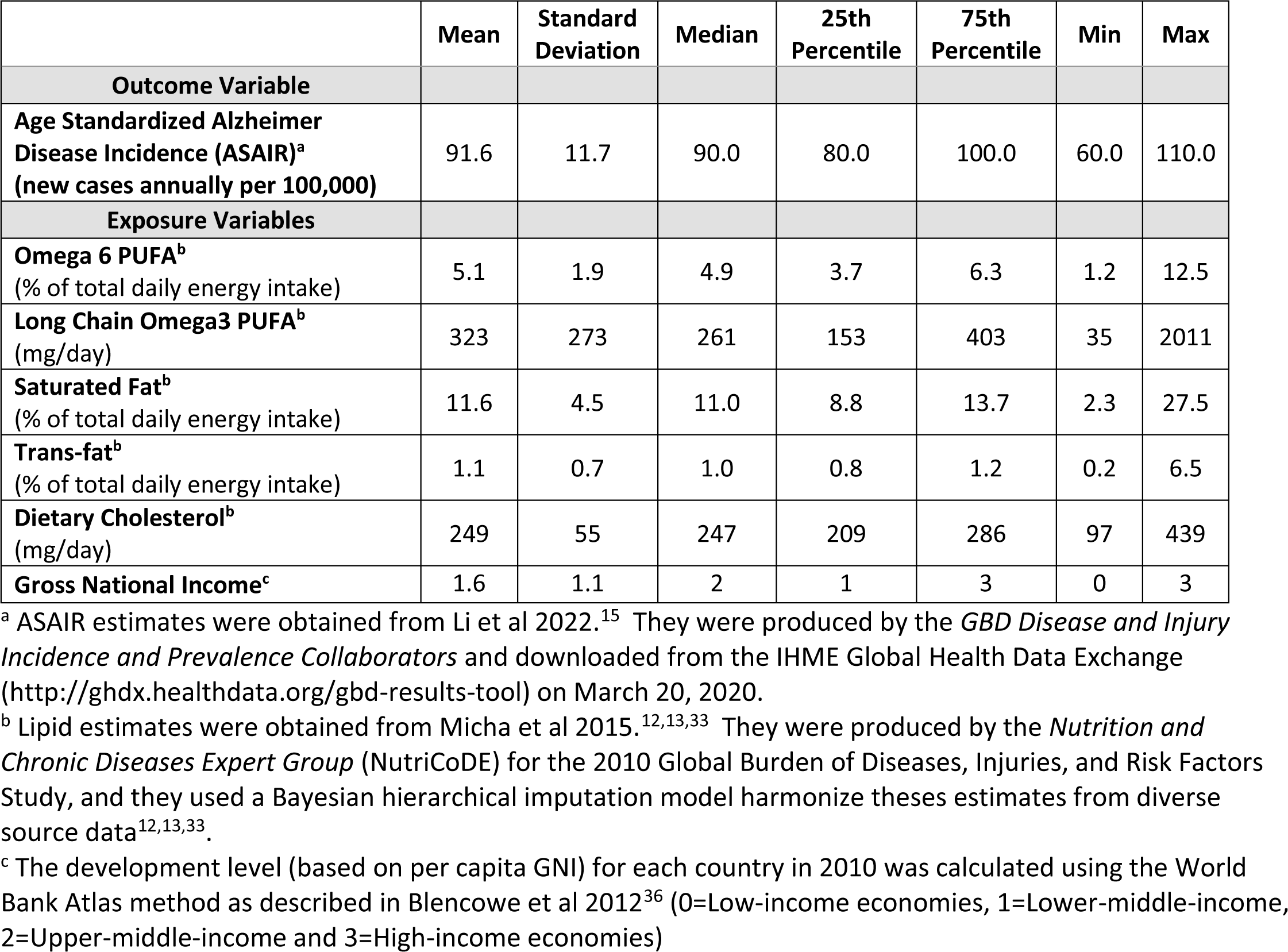
Descriptive statistics for the 183 countries.

### Cross-sectional Analyses with a Lag

Bivariate (unadjusted) free-knot penalized spline regression analyses revealed a linear relationship between country-level ASAIR and both Omega6 PUFA and Omega3 PUFA (Figure 1: A and B), but non-linearity was detected for the remaining three lipids: saturated fat, trans-fat, and cholesterol intake (Figure 1: C, D, and E). ASAIR increased with saturated fat up to ∼10% of total energy intake, and with trans-fat up to ∼1.5% total energy intake. ASAIR also increased with dietary cholesterol intake up to ∼250 mg/day. Below these thresholds the relationships appeared linear, while above these thresholds data was sparse. Thus, the slopes in the high intake regions of the exposure distribution could not be estimated with confidence, although the plots generally showed slopes near zero (Figure 1).

**Figure 1.**
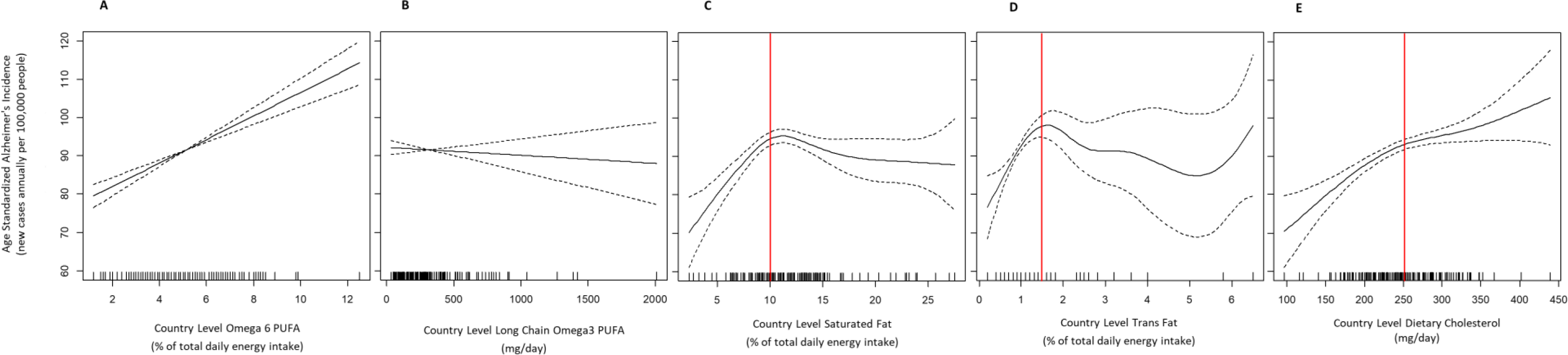
Free-knot penalized splines showing the relationship between country-level lipid intakes and age standardized incidence of Alzheimer Disease and other dementias. Incidence is on the y-axis and the y axis range is the same for all 5 plots, for ease of comparison. Each plot depicts the penalized regression spline for a distinct class of country-level lipid data: (A) Omega6 PUFA (B) Long Chain Omega3 PUFA (C) Saturated Fat (D) Trans-fat and (E) Dietary Cholesterol intake. The dashed lines depict 95% CIs and the hash marks along the x-axis depict data density. Estimates are imprecise in regions of low data density. Nonlinear relationships were detected by the GCV process for Saturated Fat, Trans-fat, Dietary Cholesterol (panels C-E). Apparent thresholds are indicated with a vertical red line.

A multivariable linear regression model adjusted for all other lipids and country development level (per capita GNI), revealed a positive association between Omega6 PUFA and ASAIR (Table 2; β = 2.44; 95%CI: 1.70, 3.19; p=1.38×10^-9^). For Omega3 PUFA there was a subtle negative trend, but the relationship was not significant (β = -0.004; 95%CI: -0.010, 0.001; p=1.11×10^-1^). Because the remaining lipids were modeled with nonlinear splines, it was not appropriate to estimate a single slope across the full exposure distribution for these lipids. However, as noted above, the remaining lipid-ASAIR relationships appeared linear below lipid-specific thresholds. Thus, for the three remaining lipids, we stratified by that lipid’s specific threshold, and reran the multivariable model using a linear term for that lipid. This allowed us to estimate slopes (βs) within the linear regions of the relationship. Below the threshold of 10% of total energy intake, we found a significant association between saturated fat and ASAIR (β = 2.67; 95%CI: 1.23, 4.11; p=5.71×10^-4^). This relationship was not evidence above the threshold (β = 0.04; 95%CI: -0.44, 0.53; p=8.62×10^-1^). Similarly, trans-fat intake was significantly associated with ASAIR below the threshold of 1.5% total energy (β = 7.35; 95%CI: 0.79, 13.90; p=2.96×10^-2^), but not above the threshold (β = 0.70; 95%CI: -1.89, 3.29; p=6.03×10^-1^). Finally, the relationship between cholesterol intake and ASAIR was significant below the threshold of 250 mg/day (β = 0.15; 95%CI: 0.07, 0.23; p=5.98×10^-4^), but not above it (β = 0.02; 95%CI: -0.03, 0.07; p=3.61×10^-1^).

**Table 2:** Results from regression models of age standardized Alzheimer Disease incidence on the country-level.

| | Change in Age Standardized<br>Alzheimer Disease Incidence<br>associated with a 1 unit increase<br>in country-level lipid ($\beta$) | 95% CI | p value |
| --- | --- | --- | --- |
| **Omega6 PUFA (% of energy intake) <sup>a</sup>** | 2.44 | 1.70, 3.19 | $1.38 \times 10^{-9}$ |
| **Long Chain Omega3 PUFA (mg/day) <sup>a</sup>** | -0.004 | -0.010, 0.001 | $1.11 \times 10^{-1}$ |
| **Gross National Income <sup>a</sup>** | 0.29 | -1.34 1.92 | $7.28 \times 10^{-1}$ |
| **Saturated Fat (% of energy intake) <sup>b</sup>** |  |  |  |
| Below the threshold of 10 | 2.67 | 1.23, 4.11 | $5.71 \times 10^{-4}$ |
| Above the threshold of 10 | 0.04 | -0.44, 0.53 | $8.62 \times 10^{-1}$ |
| **Trans-fat (% of energy intake) <sup>c</sup>** |  |  |  |
| Below the threshold of 1.5 | 7.35 | 0.79, 13.90 | $2.96 \times 10^{-2}$ |
| Above the threshold of 1.5 | 0.70 | -1.89, 3.29 | $6.03 \times 10^{-1}$ |
| **Dietary Cholesterol (mg/day) <sup>d</sup>** |  |  |  |
| Below the threshold of 250 | 0.15 | 0.07, 0.23 | $5.98 \times 10^{-4}$ |
| Above the threshold of 250 | 0.02 | -0.03, 0.07 | $3.61 \times 10^{-1}$ |

a Multivariable model with Omega6 PUFA, Long Chain Omega3 PUFA (mg/day) and Gross National Income modeled with linear terms, and Saturated Fat, Dietary Cholesterol, and Trans-fat were modeled with penalized splines.
b The same model as in a, but now with a linear term for Saturated Fat, and evaluated in two separate strata (above and below the threshold of 10% Saturated fat by total energy intake)
c The same model as in a, but now with a linear term for Trans-fat, and evaluated in two separate strata (above and below the threshold of 1.5% Trans-fat by total energy intake)
d The same model as in a, but now with a linear term for Dietary Cholesterol, and evaluated in two separate strata (above and below the threshold of 250 mg/day of Dietary Cholesterol)

### Longitudinal Analyses

The longitudinal within-country changes in lipid intakes between 1990 and 2010 were small (Table 3), but the unadjusted regression model validated the association between Omega6-PUFA and ASAIR (Table 4; β = 0.0341; 95%CI: 0.0154, 0.0529; p=4.70×10^-4^). Unadjusted regression models for the other lipids yielded findings that were generally consistent with the cross-sectional findings, but none of the associations reached significance (Table 4). The longitudinal relationship between Omega6-PUFA and ASAIR remained significant upon multivariable adjustment (Table 4; β = 0.0375; 95%CI: 0.0164, 0.0586; p=6.50×10^-4^).

**Table 3:** Longitudinal changes in lipid intakes and age standardized Alzheimer Disease incidence in the 183 countries.

|  | Mean | Standard Deviation | Median | 25th Percentile | 75th Percentile | Min | Max |
| --- | --- | --- | --- | --- | --- | --- | --- |
| **Outcome Variable** |  |  |  |  |  |  |  |
| **Percent Change in Annual Age Standardized Alzheimer Disease Incidence from 1990 to 2019<sup>a</sup>** | 0.012 | 0.111 | 0.020 | -0.040 | 0.070 | -0.420 | 0.580 |
| **Exposure Variables** |  |  |  |  |  |  |  |
| **Percent Change in Omega 6 PUFA from 1990 to 2010<sup>b</sup>** | 0.42 | 0.83 | 0.30 | -0.10 | 0.70 | -2.00 | 3.40 |
| **Percent Change in Long Chain Omega3 PUFA from 1990 to 2010<sup>b</sup>** | 58 | 79 | 41 | 9 | 94 | -193 | 400 |
| **Percent Change in Saturated Fat from 1990 to 2010<sup>b</sup>** | 0.05 | 1.32 | 0.00 | -0.60 | 0.50 | -4.40 | 4.60 |
| **Percent Change in Trans-fat from 1990 to 2010<sup>b</sup>** | 0.07 | 0.21 | 0.00 | 0.00 | 0.10 | -0.80 | 1.50 |
| **Percent Change in Dietary Cholesterol from 1990 to 2010<sup>b</sup>** | 3.6 | 19.9 | 2.2 | -10.3 | 15.7 | -57.0 | 57.8 |
| **Change in Gross National Income from 1990 to 2010<sup>c</sup>** | 0.38 | 0.60 | 0 | 0 | 1 | -1 | 2 |

<sup>a</sup> ASAIR estimates were obtained from Li et al 2022.<sup>15</sup> They were produced by the *GBD Disease and Injury Incidence and Prevalence Collaborators* and downloaded from the IHME Global Health Data Exchange (http://ghdx.healthdata.org/gbd-results-tool) on March 20, 2020.
<sup>b</sup> Lipid estimates were obtained from Micha et al 2015.<sup>12,13,33</sup> They were produced by the *Nutrition and Chronic Diseases Expert Group* (NutriCoDE) for the 2010 Global Burden of Diseases, Injuries, and Risk Factors Study, and they used a Bayesian hierarchical imputation model harmonize these estimates from diverse source data<sup>12,13,33</sup>.
<sup>c</sup> This variable was coded as the country development level change (based on per capita GNI) from 1990 to 2010. Development levels were not available for 27 countries in 1990, but we were able to obtain substitute country development baseline values for 24 of these countries by using an estimate from 1991 or 1992.<sup>37</sup> Thus, only Montenegro, Serbia, and Timor-Leste were missing country development level change data.

**Table 4:** Results from linear regression models of percent change in annual age standardized Alzheimer Disease Incidence from 1990 to 2019.

| | Percent Change in Annual Age Standardized Alzheimer Disease Incidence from 1990 to 2019 associated with a 1% increase in country-level lipid intake ($\beta$) | 95% CI | p value |
| --- | --- | --- | --- |
| **Unadjusted Models** |  |  |  |
| **Percent Change in Omega 6 PUFA from 1990 to 2010** | $3.41 \times 10^{-02}$ | $1.54 \times 10^{-02}$, $5.29 \times 10^{-02}$ | $4.70 \times 10^{-04}$ |
| **Percent Change in Long Chain Omega3 PUFA from 1990 to 2010** | $9.63 \times 10^{-5}$ | $-1.08 \times 10^{-4}$, $3.01 \times 10^{-4}$ | $3.58 \times 10^{-1}$ |
| **Percent Change in Saturated Fat from 1990 to 2010** |  |  |  |
| Below the threshold of 10 | $4.38 \times 10^{-3}$ | $-1.83 \times 10^{-2}$, $2.70 \times 10^{-2}$ | $7.06 \times 10^{-1}$ |
| Above the threshold of 10 | $-2.39 \times 10^{-3}$ | $-1.71 \times 10^{-2}$, $1.23 \times 10^{-2}$ | $7.51 \times 10^{-1}$ |
| **Percent Change in Trans-fat from 1990 to 2010** |  |  |  |
| Below the threshold of 1.5 | $1.37 \times 10^{-1}$ | $-2.43 \times 10^{-2}$, $2.98 \times 10^{-1}$ | $9.82 \times 10^{-2}$ |
| Above the threshold of 1.5 | $2.28 \times 10^{-2}$ | $-6.33 \times 10^{-2}$, $1.09 \times 10^{-1}$ | $6.08 \times 10^{-1}$ |
| **Percent Change in Dietary Cholesterol** |  |  |  |
| Below the threshold of 250 | $9.23 \times 10^{-4}$ | $-2.44 \times 10^{-4}$, $2.09 \times 10^{-3}$ | $1.24 \times 10^{-1}$ |
| Above the threshold of 250 | $2.14 \times 10^{-5}$ | $-1.16 \times 10^{-3}$, $1.20 \times 10^{-3}$ | $9.72 \times 10^{-1}$ |
| **Multivariable Adjusted Model<sup>a</sup>** |  |  |  |
| **Percent Change in Omega 6 PUFA from 1990 to 2010** | $3.75 \times 10^{-02}$ | $1.64 \times 10^{-02}$, $5.86 \times 10^{-02}$ | $6.50 \times 10^{-04}$ |

<sup>a</sup> Adjusted for: Percent Change in Long Chain Omega3 PUFA, Percent Change in Saturated Fat, Percent Change in Trans-fat, and Percent Change in Dietary Cholesterol, and change in Gross National Income level. To account for distinct associations on opposite sides of the threshold for saturated fat, the saturated fat variable was interacted with a dichotomous dummy variable for *change-occurred-above-or-below threshold*. Trans-fat and dietary cholesterol also had thresholds and were modeled in the same way.

## Discussion

We observed a multivariable-adjusted positive linear association between mean country-level intake of Omega6 PUFA in 2010 and ASAIR in 2019. This association was then validated with a second multivariable adjusted model using longitudinal data. Specifically, we found that the percent change in the country-level Omega6 intake from 1990 to 2010 was positively associated with the percent change ASAIR between 1990 and 2019. These findings need further validation and trials to establish if the relationship is causal. However, the validation of this pattern in longitudinal data is among the strongest corroborations that can be made in non-experimental data. We adjusted for all other lipids to reduce the risk of bias. With a sample size of 183 countries, we found strong associations in discovery and in longitudinal validation. These are exceptionally small p-values for an epidemiologic study with this sample size. Our findings remain significant even under the most conservative adjustment for multiple testing (considering both discovery and confirmatory regressions as 18 separate tests the Bonferroni adjusted threshold for α=0.05 would be p<2.8×10^-3^)

The association between Omega6 PUFA and Alzheimer Disease is consistent with prior literature in individual level human studies and animal experiments^8,16–20^. While other studies have reported an inverse association between Omega3 PUFA and AD^8,21^, here we only observed a non-significant trend in this direction. Omega3 and Omega6 PUFA share an endogenous processing pathway^22^ involving fatty acid desaturases (FADS) and elongases of very long chain fatty acids (ELOVL). Interestingly, the gene clusters that produce these enzymes are both implicated in AD ^16,23^. Thus, our findings are consistent with the hypothesis that the inflammatory endpoints of Omega6 processing (e.g. Arachidonic Acid) are more important than the Omega-3 physiology for increasing the risk of Alzheimer disease.

The magnitude of the initial association with Omega6 (β = 2.44; 95%CI: 1.70, 3.19; p=1.38×10^-9^) indicates that a one standard deviation decrease (-1.9% as a percent of total daily energy intake) could reduce ASAIR by 4.6 new cases of Alzheimer disease per 100,000 per year (2.44*1.9). For the US, which reported an ASAIR of 110 new cases per 100,000 in 2019, a 2 standard deviation decrease in mean Omega6 PUFA intake would amount to 8.4% reduction in new cases annually ([110-9.2]/110 = 91.6%). In absolute numbers this translates into ∼28,140 fewer new cases each year (8.4*[335,000,000/100,000]). We did not evaluate Alzheimer Disease prevalence in this study because prevalence is affected by survival time, which can involve factors that are not relevant to disease etiology. However, if all other factors were held constant, lower annual incidence could compound over time to reduce the prevalence and societal burden of Alzheimer Disease.

Our findings for saturated fat, trans-fat, and dietary cholesterol are also supported by the literature^2,24–27^, but the patterns were not significant in our longitudinal analyses. This is not surprising given the small magnitude of the longitudinal changes in lipid intakes that were observed within countries. Individual level studies, both cross-sectional and longitudinal, generally share the same limitation (i.e. small range of exposure). The results from our free-knot penalized spline regression models remain critical for future research as they indicate that nonlinear relationships should be considered in future inquiries of these lipids. If the relationships reported here are validated in subsequent work, then reductions in saturated fat, trans-fat, and dietary cholesterol may further reduce country-level ASAIR. For countries below the saturated fat threshold of 10% of total daily energy intake, a two standard deviation decrease in saturated fat intake would correspond to a reduction in ASAIR by 24.0 new cases of Alzheimer Disease per 100,000 per year (2.67*9.0). Similar calculations for trans-fat (7.35*1.4) and dietary cholesterol (0.15*110) would predict 10.3 and 16.5 fewer new cases per 100,000 per year, respectively. In 2019 the US had a mean saturated fat intake of 11.8% of total daily energy intake, a mean trans-fat intake of 2.8% of total daily energy intake, and a mean dietary cholesterol of 296 mg/day. A two standard deviation decrease for each of these fats would lower the US mean intakes below the identified intake thresholds (10% of total daily energy intake for saturated fat, 1.5% of total daily energy intake for trans-fat, and 250 mg/day for dietary cholesterol). Such a decrease would result in 19.2 fewer new Alzheimer Disease cases per 100,000 per year for saturated fat reductions (2.67*7.2), 0.7 fewer new cases per 100,000 per year for trans-fat reductions (7.35*0.1), and 9.6 fewer new cases per 100,000 per year for dietary cholesterol reductions (0.15*64). When added to the Omega6 PUFA findings, a two standard deviation decrease in Omega6, Saturated fat, trans-fat, and dietary cholesterol could potentially yield a net reduction of 38.7 new cases per 100,000 per year in the US. This would amount to a 35.2% reduction in new cases of Alzheimer Disease in the US each year. ([110-38.7]/110 = 64.8%). In absolute numbers this translates into ∼129,645 fewer new cases each year in the US (38.7*[335,000,000/100,000]). Again, the saturated fat, trans-fat, and dietary cholesterol findings still need to be corroborated on the country level, but there is substantial motivation to conduct follow-up studies because of the size of the putative benefits.

There are limitations to our work. First, we note that our country-level analyses depend on the accurate estimation of mean lipid intakes and ASAIR. These estimates come from peer reviewed publications and extensive care was made to account for potential biases. Second, we note that we had limited covariates available for confounding adjustment and this is a common issue in country-level analyses. Nevertheless, we emphasize that the findings in the longitudinal analyses intrinsically account for confounding factors that are stable in time. In other words, confounding by genetic and environmental factors that are stable within the time course of the analysis, are controlled for by study design. Additionally, we adjusted both our cross-sectional and longitudinal analyses for country development level. This can reduce residual bias from factors that are correlated with country level economic improvement. Perhaps most importantly, the Omega6 PUFA results were validated in the longitudinal models despite limited power (and the other findings revealed supportive trends). A third limitation is that the time lags do not perfectly align between the discovery and validation analyses. We would have preferred to have access to changes in ASAIR between 1999 and 2019 instead of 1990 to 2019. However, this is a minor point and the expected random error would likely bias toward the null. Finally, we note that while these findings are very useful for country-level inference and intervention, they are not directly applicable to individual level inference (the ecological fallacy). Therefore, although our results are compelling at the country level, the individual level will require additional study.

Overall, these findings may provide an approach for reducing ASAIR through food system interventions. If trials confirm our findings, then we can hone the prevention strategies that governments need as populations around the globe get older.^28^ Importantly, this work stands on vast convergent evidence^29^ implicating lipid dysfunction in Alzheimer disease etiology.^1–9^ It is now becoming clear that lipid abnormalities are causal factors^30^ in Alzheimer disease development, and though lipids have not always been the center of focus for Alzheimer researchers, the genetics have been consistently pointing in this direction for over 30 years.^1,7^ Perhaps most importantly, if achieved, diet-based interventions could have large impact on Alzheimer disease incidence. While determining safe, optimal changes to country level lipid intakes needs some additional study, the benefits might extend beyond ASAIR, as excess Omega6 PUFA in our food systems is linked to many other serious and common chronic diseases.^31,32^ The potential ASAIR benefits will vary by country, but based on our estimates for the US, a moderate decrease in Omega6 PUFA intake could reduce ASAIR by 8%. Additional decreases in saturated fat, trans-fat, and dietary cholesterol might yield total ASAIR reductions of 35%. Our findings are promising as they indicate a rather straightforward path to reducing AD burden.

## Online Methods

Previously published reports were used to obtain mean intake for 5 lipids (Omega6 polyunsaturated fatty acids (Omega6 PUFA), Long Chain Omega3 polyunsaturated fatty acids (Omega3 PUFA), Saturated fat, Trans-fat, and Dietary Cholesterol)^12,13^ and ASAIR^15^ for 184 countries. One country, the Maldives, was eliminated from the analyses as it was an extreme outlier with respect to Omega3 PUFA (>9 standard deviations above the global mean). The lipid estimates were produced by the Nutrition and Chronic Diseases Expert Group (NutriCoDE) for the 2010 Global Burden of Diseases, Injuries, and Risk Factors Study, and they used a Bayesian hierarchical imputation model to harmonize these estimates from diverse source data^12,13,33^. ASAIR estimates were produced by the GBD Disease and Injury Incidence and Prevalence Collaborators and downloaded from the IHME Global Health Data Exchange (http://ghdx.healthdata.org/gbd-results-tool) on March 20, 2020^15^ Incidence rates in this study are for Alzheimer disease and other dementias (national autopsy programs are not available for diagnostic confirmation at scale). Age standardization was conducted to account for country-level differences in population age distributions and age of dementia onset. Lipid intake data was from 2010 and ASAIR values were from 2019. This 9-year time-lag aligns with optimal lags estimated for environmental factors in Alzheimer Disease epidemiology.^34^ Omega3 PUFA estimates were adjusted to account for endogenous conversion as previously described^35^.

### Cross-sectional Analyses with a Lag

To assess the shape of the 5 lipid-ASAIR relationships, we implemented free-knot penalized regression splines using MGCV in R.^14^ The three lipid intakes that demonstrated non-linear relationships with ASAIR in these bivariate regression spline models were specified in the subsequent multivariable-adjusted model with spline terms. Limited covariate data was available for confounding adjustments, but this model was adjusted for all other lipids and country development level based on per capita Gross National Income (GNI)^36^. The per capita GNI for each country in 2010 was calculated using the World Bank Atlas method as described in Blencowe et al 2012^36^ (0=Low-income economies, 1=Lower-middle-income, 2=Upper-middle-income and 3=High-income economies). Since the three nonlinear relationships appeared to be linear on either side of a threshold in the bivariate spline models, we also used the thresholds to create 2 strata for each of these lipids (strata within which the lipid-ASAIR relationship appears linear). We then specified linear terms for these lipids and reran the multivariable-adjusted regression model in the two strata separately (to obtain βs for the linear regions of the lipid-ASAIR relationship).

### Longitudinal Analyses

Finally, we compared the percent change in lipid intakes from 1990 to 2010^12,13^ to the percent change in ASAIR between 1990 and 2019^15^ with linear regression. We first assessed unadjusted linear regression models within the linear regions of the relationship identified in the analyses described above. Any lipid that yielded significant findings was then evaluated in a multivariable-adjusted model that was adjusted for all other lipids. To identify distinct associations on opposite sides of the thresholds for saturated fat, trans-fat, and dietary cholesterol, we tested for interaction with a dichotomous dummy variable for change-occurred-above-or-below threshold. Finally, we adjusted for changes in country development level based on per capita GNI. This variable was coded as the development level change from 1990 to 2010. Development levels were not available for 27 countries in 1990, but we were able to obtain substitute baseline values for 24 of these countries by using development estimates from 1991 or 1992.^37^ Thus, only Montenegro, Serbia, and Timor-Leste were excluded due to missing baseline development data in any of these years. Note that the lipid intake information is the percent change in the country-level intake between 1990 and 2010, and the incidence data is percent change in the annual age standardized incidence between 1990 and 2019. Since these units differ from the prior analyses, these data can be used to corroborate association directions, but the association magnitudes are not directly comparable. Descriptive statistics were obtained using SAS 9.4 (Cary, NC) and all other analyses were conducted with R 4.2.1.

## Data Availability

The country-level data that we used is available from the previously published source papers, as specified in the manuscript.

## Acknowledgements

The authors would like to thank the many people who contributed to the country-level estimates that were used in these analyses.

## Consent statement

NA – This analysis used country-level data from previously published papers.

## Research in Context

### Evidence before this study

Convergent evidence from genetics, neuropathology, laboratory experiments, and epidemiology indicates that lipids play a central role in the development of Alzheimer Disease (AD). However, the relative importance of different lipids remains unclear, and individual-level epidemiologic studies are often not well suited to address this question. Large cohort studies rarely have precise dietary information on specific lipid intakes over decades, and even where these data are available, the lipid intakes among individuals do not typically vary enough within a study population to discern patterns of association. Additionally, multiple categories of lipids are not usually available in one dataset, so their relative associations cannot be compared. Lastly, the potential for nonlinear relationships is rarely considered. These caveats often lead to equivocal conclusions upon meta-analysis. However, each of these issues is addressable with analyses on the country-level.

### Added value of this study

Here we obtained country-level data on five lipid intakes, and compared them to age-standardized Alzheimer Disease Incidence rates (ASAIR) using free-knot penalized spline regression and linear regression. We found that Omega6 Polyunsaturated-Fatty-Acid (PUFA) intake exhibited a linear and significant relationship with ASAIR and it was validated in a follow-up longitudinal analysis. Additionally, we found non-linear relationships between saturated-fat, trans-fat, dietary-cholesterol and ASAIR. For each of these, ASAIR increased with lipid intake up to a threshold although these relationships were not statistically significant in the longitudinal analysis.

### Implications of all the available evidence

When combined with the pre-existing evidence, these analyses indicate that decreasing Omega6-PUFA consumption on the country-level may have substantial benefits in reducing AD rates. Further research is needed on all these lipids, but country level reductions in saturated-fat, trans-fat, dietary-cholesterol intake, hold similar promise for reducing the country level burden of dementia. When considered in aggregate, reductions in the mean intake of four lipids may yield significant reductions in AD incidence. Individual level studies must be conducted to determine the impact of lipid intakes on specific people.

## Highlights

Diverse convergent evidence indicates that lipids play a role in Alzheimer Disease.

Individual-level studies are often not well suited to study lipid consumption.

Incidence of Alzheimer Disease and other dementias increased with mean country-level Omega6 PUFA intake

It also increased with saturated-fat, trans-fat, and cholesterol up to a threshold.

The country-level Omega6 PUFA finding was validated in longitudinal analyses.

